# Operational TBE incidence forecasts for Austria, Germany, and Switzerland 2019–2021

**DOI:** 10.1101/2020.07.09.20149492

**Authors:** Franz Rubel, Katharina Brugger

## Abstract

In spring 2019, forecasts of the incidence of tick-borne encephalitis (TBE) for the next two years, i.e. 2019 and 2020, were made for the first time. For this purpose, negative binomial regression models with 4–5 predictors were fitted to the time series of annual human TBE incidences from Austria, Germany and Switzerland. The most important predictor for TBE incidences is the fructification index of the European beech (*Fagus sylvatica*) 2 years prior as a proxi for the intensity of the TBE virus transmission cycle. These forecasts were repeated in spring 2020 after the updated predictors and the confirmed TBE cases for 2019 became available. Forecasting TBE incidences for 2020 and 2021 results in 156±19 and 131±23 TBE cases for Austria, 663±95 and 543±112 TBE cases for Germany as well as 472±56 and 350±62 TBE cases for Switzerland. The newly implemented operational TBE forecasts will be verified every year with confirmed TBE cases. An initial verification for 2019 demonstrates the high reliability of the forecasts.

## 1. Introduction

Tick-borne encephalitis (TBE) is a viral tick-borne zoonosis that is endemic in wide areas of Eurasia from eastern France to Russia and China (Dobler et al., 2019). Although of great public interest, an operational TBE forecast is only available for Russia (Noskov et al., 2017). It is based on a trend analysis and published annually in a scientific journal, most recently by Nikitin et al. (2020).

A more sophisticated method is used here, which takes into account the trend as well as the low- and high-frequency oscillations in the human TBE incidence series. The frequency spectra of various European TBE time series were first analyzed by Zeman (2017a), who described 10 years and 2–3 years dominating periods of the oscillations. He demonstrated that a superposition of several distinct periods, that are relatively stable and synchronous over Central Europe, can be extrapolated into the future (Zeman, 2017b). The advantage of these harmonic regression models is that they do not require biological or climatological predictors. The disadvantage is that natural deviations from the two dominating periods of the oscillations cannot be predicted. Therefore, biological and climatological predictors have recently been proposed to explain the oscillations in the TBE incidence series (Rubel et al., 2020). Accordingly, the high frequency oscillations, i.e. those oscillations responsible for the next years TBE incidences, are well described by the fructification index of the European beech (*Fagus sylvatica*). This applies at least to the countries of Austria, Germany and Switzerland, where the prevailing climate at the lower altitudes is classified according to Köppen-Geiger as Cfb-climate (Rubel et al., 2017). The Cfb-climate is also referred to as a beech climate, after the tree species dominating in natural forests. Beech fructification is a predictor of the intensity of the natural TBE virus transmission cycle between small mammals (rodents) and its main vector *Ixodes ricinus*, with a high beech fructification index increasing the population density of rodents and of *I. ricinus* larvae one year thereafter. One more year later, significantly higher densities of questing *I. ricinus* nymphs are responsible for the more frequent transmission of the TBE virus to humans (Brugger et al., 2018). The beech fructification can thus be used to predict the TBE incidences of the next two years. Particularly, years with full fructification of beech (mast seeding) are responsible for several high peaks in the TBE time series. This was demonstrated by the recently developed TBE models for Austria, Germany and Switzerland, which were used to forecast the national TBE cases for 2019 and 2020 (Rubel and Brugger, 2020). These TBE models are now used in an operational forecast scheme including verification with independent data.

## 2. Material and methods

Negative binomial regression models are applied that were originally developed to explain the oscillations in the Austrian long-term TBE incidence series (Rubel et al., 2020). They use two demographic parameters, the vaccination coverage, a climate index, and a beach fructification index as predictors. Details concerning the TBE models and its predictors were described by Rubel and Brugger (2020), who extended the models to forecast Austrian, German and Swiss TBE incidences for 2019 and 2020. For the first operational application, the forecasts of national TBE incidences for 2020 and 2021 and the verification of the forecasts for 2019, all TBE incidence series and predictors have been updated with new data provided by official sources in spring 2019. The availability of all data is also described in detail by Rubel and Brugger (2020). An exception is the beech fructification index after Konnert et al. (2016). There is no official publicly available database for this most important TBE predictor. The updated values of the beech fructification index 2017-2019, which are used for the TBE incidences forecasts 2019–2021, were provided by Anneliese Nitzinger (Bavarian Office for Forest Genetics) on personal request. Accordingly, a sporadic occurrence of beech fructification, but not noticeable at first sight, was observed for 2017 and 2019. In 2018, however, full beech fructification was observed, also known as mast seeding. The following updated values of the fructification index two years prior written in the nomenclature of Rubel and Brugger (2020) are used: *F*_*year*−2_ = 1 for 2019, *F*_*year*−2_ = 3 for 2020 and *F*_*year*−2_ = 1 for 2021.

The goodness-of-fit of the calibrated TBE forecast models is verified by the root-mean-square error RMSE, the explained variance R^2^, and the adjusted variance 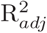. Additionally, a scattergram shows the correlation R and the corresponding significance level (p-value). The absolute errors of the mean as well as the mean ± standard deviation forecasts are given for the verification with independent data (currently only available for 2019).

## 3. Results

The first result of this work is the forecast of annual human TBE incidences for 2020 and 2021 as calculated in spring 2020 (Fig. 1). The time series plots depict the observed TBE incidences (grey bars) overlaid with the prediction of the calibration period 1991–2019 (red lines) and the forecast period 2020–2021 (red dotted lines). Following TBE incidences were forecasted for 2020 and 2021: 156±19 and 131±23 TBE cases for Austria, 663±95 and 543±112 TBE cases for Germany as well as 472±56 and 350±62 TBE cases for Switzerland.

**Figure 1:**
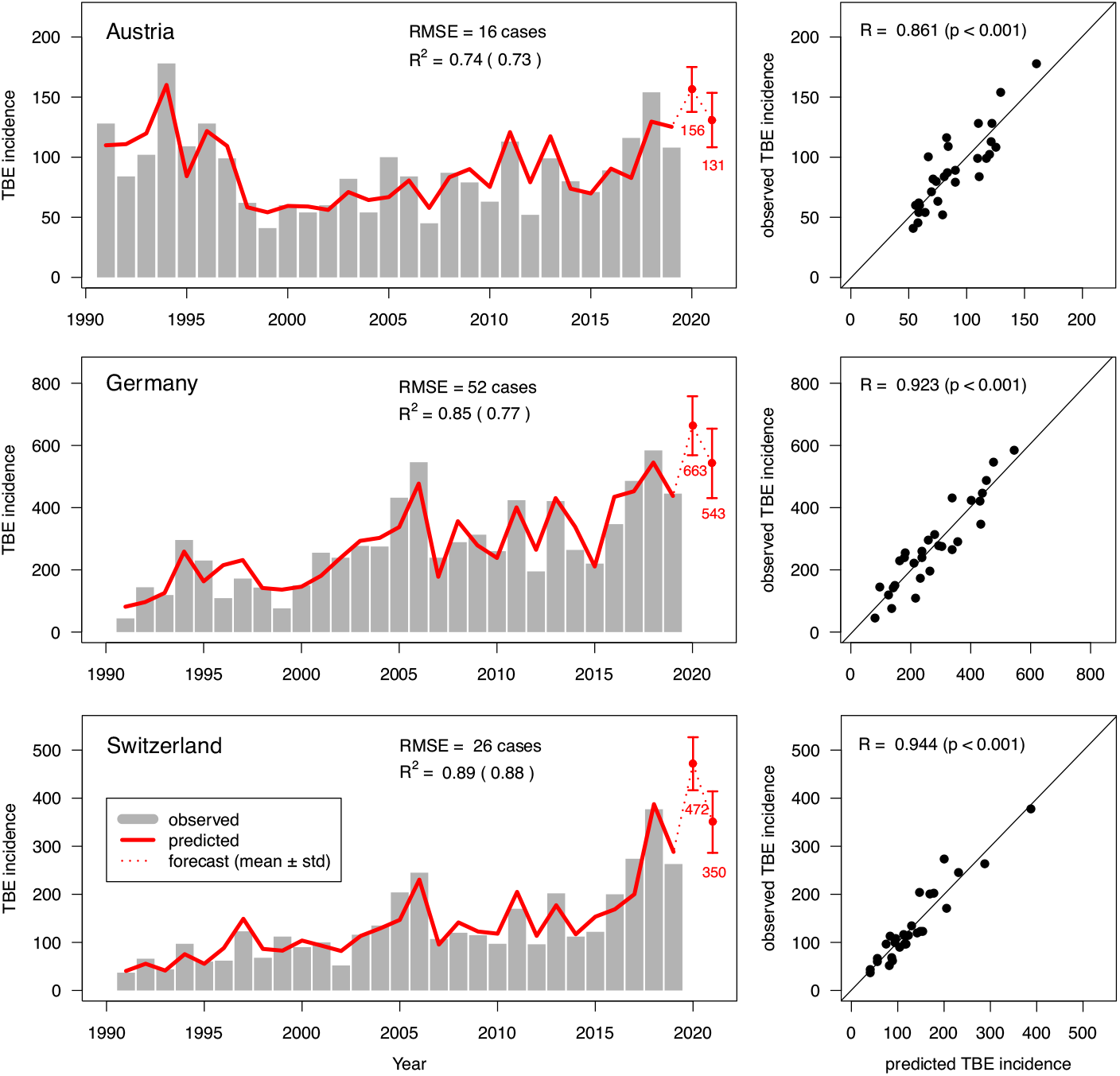
Austrian, German, and Swiss tick-borne encephalitis (TBE) incidences. Left: Time series of observed (grey bars), predicted for the calibration period 1991–2019 (red line) and predicted for the forecast period 2020–2021 (red dotted line). Right: Scatterplots predicted vs. observed TBE incidence. Verification measures include root-mean-square error (RMSE), explained variance R^2^ 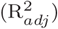, and the correlation coefficient R (p-value) shown in the scatterplots.

The second result is an operational forecast scheme with verification based on independent observations (Table 1). According to the TBE forecasts for 2019 and 2020, as of spring 2019, forecasts are currently available for three years. There are even two different forecasts for 2020 that can be compared. These are the forecasts as of spring 2019 for the upcoming year 2020 (year +2) and the forecasts as of spring 2020 for the entire year 2020 (year +1). The forecasts for 2020 should be able to be verified with the reported TBE cases available in spring 2021. Then the year + 1 forecasts should be more accurate than the year + 2 forecasts. Currently only the forecasts for 2019 can be verified. These forecasts are systematically too low by 16 TBE cases for Austria, 28 TBE cases for Germany and 8 TBE cases for Switzerland. However, only the forecast for Austria is slightly outside the forecast interval of 92±12 cases (108 cases observed, error=104-108=-4 cases). The forecasts can therefore be classified as reliable.

**Table 1:** Operational forecast and verification scheme for Austrian, German, and Swiss human tick-borne encephalitis incidences comprising year+2 and year+1 forecasts, observations, and errors of mean (mean ± standard deviation). Data available in spring 2021, 2022 and 2023 are marked by *, ** and ***.

| Country | Year | Forecasts |  | Observations | Errors |  |
| --- | --- | --- | --- | --- | --- | --- |
|  |  | +2 | +1 | 0 | +1 | +2 |
| Austria | 2019 | | 92 ± 12 | 108 | 16 (-4) | * |
| | 2020 | 142 ± 26 | 156 ± 19 | * | * | ** |
| | 2021 | 131 ± 23 | * | ** | ** | *** |
| Germany | 2019 | | 417 ± 71 | 445 | 28 (0) | * |
| | 2020 | 670 ± 168 | 663 ± 95 | * | * | ** |
| | 2021 | 543 ± 112 | * | ** | ** | *** |
| Switzerland | 2019 | | 235 ± 30 | 263 | 8 (0) | * |
| | 2020 | 465 ± 91 | 472 ± 56 | * | * | ** |
| | 2021 | 350 ± 62 | * | ** | ** | *** |

## 4. Discussion

Tick-borne encephalitis is well-suited for prediction. The necessary causal relationships with the natural TBE virus transmission cycle between rodents and ticks have been known for a long time. This applies to the relationships between the fructification of forest trees (masting in trees) and the density of rodents (Ostfeld et al., 1996; Koenig and Knops, 2005; Clement et al., 2010), but also for the one year time-lag between high rodent density and high human TBE incidences (Balashov, 2012). If long time series of rodent density are available, these can also be used as predictors for TBE incidences forecasts for the next year (Tkadlec et al., 2019). Since rodent density affects the density of their predators, such as red foxes (*Vulpes vulpes*), these are also suitable predictors for human TBE incidences (Haemig et al., 2011). However, the latter must be estimated from hunting indices, which is associated with uncertainties.

A limiting factor of the forecasts of the next 2 years’ TBE incidences is the availability of latest TBE case numbers and demographic data provided by the national authorities. The real-time availability of the beech fructification index is also problematic. These data are usually updated in the spring of the following year, which is why the forecasts can only be made afterwards. Although the first results are encouraging, a final statistical verification of the operational TBE incidences forecasts can only be made after they have been available for at least 7–10 years.

## Data Availability

Only publicly accessible data was used

